# Systematic review and meta-analysis of the accuracy of McIsaac and Centor score in patients presenting to secondary care with Pharyngitis

**DOI:** 10.1101/2023.02.22.23286307

**Authors:** Atchchuthan Kanagasabai, Callum Evans, Hayley E Jones, Alastair D Hay, Sarah Dawson, Jelena Savović, Martha M C Elwenspoek

**Affiliations:** Population Health Sciences, Bristol Medical School, University of Bristol, BS8 2PS Bristol, UK; Centre for Academic Primary Care, Bristol Medical School, University of Bristol, Bristol; The National Institute for Health Research Applied Research Collaboration West (NIHR ARC West), University Hospitals Bristol and Weston NHS Foundation Trust, BS1 2NT Bristol, UK

## Abstract

**Background:** Centor and modified Centor (McIsaac) scores are clinical prediction rules used to diagnose group A streptococcus infection in patients with pharyngitis. They aim to identify the patients most likely to benefit from antibiotic treatment and reduce unnecessary antibiotic prescribing.

**Objectives:** To systematically review the literature on the diagnostic accuracy of McIsaac and Centor, and produce pooled estimates of accuracy at each score threshold, in patients presenting with acute pharyngitis to secondary care.

**Data sources:** MEDLINE, Embase and Web of science were searched from inception to June 2021.

**Eligibility criteria:** Studies that included patients who presented with acute pharyngitis to hospital emergency departments and outpatient clinics, reported McIsaac or Centor scores, and used throat cultures and/or rapid antigen detection tests as the reference standard.

**Review methods:** The review protocol was registered on PROSPERO (CRD42021267413). Study selection was performed by two reviewers independently and risk of bias was assessed using the QUADAS-2 tool. Sensitivities and specificities of McIsaac and Centor scores were pooled at each threshold using bivariate random effects meta-analysis.

**Results:** The McIsaac score had higher estimated sensitivity and lower specificity relative to Centor scores at equivalent thresholds, but with wide and overlapping confidence regions. Using either score as a triage to rapid antigen detection tests (RADT) to decide antibiotic treatment would reduce antibiotic prescription to non-GAS pharyngitis patients relative to RADT test for everyone, but also reduce antibiotic prescription to GAS patients.

**Conclusion:** Our findings suggest that high thresholds of either score excludes a proportion of true positive patients from potentially beneficial treatment. The use of a low threshold before a RADT test would reduce antibiotic prescription relative to prescribing based on score only but the economics and clinical effectiveness of this combination strategy needs assessment. We recommend continued use of existing antibiotic prescribing guidelines and patient safety netting.

## Introduction

A prospective family-based cohort study of 202 families, 853 individuals, found that 16% of adults and 41% of children reported pharyngitis over one year(1). Roughly 50% of pharyngitis cases are in patients aged 5 to 24 (2). Pharyngitis is typically self-limiting and does not require medical treatment. However, between 5% and 30% of pharyngitis cases are caused by Group A streptococcus (GAS)(3), which can lead to serious suppurative and non-suppurative sequela such as otitis media, sinusitis, acute rheumatic fever, and scarlet fever (4). GAS pharyngitis is more prevalent in children (0.4 cases per person-year) than in adults (0.15 cases per person-years) (5). Prevalence of GAS pharyngitis in patients self-presenting to healthcare providers is reported as 24.1%, the same meta-analysis reports asymptomatic carriage rate across all ages as 7.0% (6). Children’s asymptomatic carriage rate (8-15.9%) was higher than adults (2.8%), although children under 5 had comparable asymptomatic carriage rate to adult (6-8).

Antibiotics are prescribed for GAS pharyngitis to prevent serious complications. Evidence suggests that antibiotics can reduce headache and sore throat symptoms at day three and sore throat symptoms at one week, incidence of acute otitis media within 14 days and rheumatic fever and quinsy within two months compared to control groups given placebo or no treatment(9).

Centor and McIsaac scores have been developed to predict the risk of GAS pharyngitis in patients presenting with pharyngitis. Centor score (range 0 to 4) is calculated from a set of four clinical parameters each scoring 1 point: tonsillar exudate, tender anterior cervical lymphadenopathy or lymphadenitis, a history of fever (over 38°C), and absence of cough (10). The McIsaac score (range -1 to 5), also called the modified Centor score, adds age as an additional criterion adding 1 point for patients aged 3-14 and subtracting 1 point for patients aged 45 or more (11).

UK guidelines recommend the use of Centor scores to stratify patients into who should be managed at home (Centor 0-2) and patients who require further investigation and potentially antibiotic treatment (Centor 3-4) (12). German (13) and Danish (14) guidelines recommend that patients with a McIsaac scores of ≥3 and ≥2, respectively, should be given a rapid antigen detection tests (RADT) and treated with antibiotics if the RADT is positive. Throat culture remains the most accurate test for confirming GAS pharyngitis in patients but requires 48-72 hours to return results. The use of much quicker RADT has increased following improvements to their sensitivity and specificity and some guidelines now recommend their use as an alternative to or before throat cultures (15, 16). A meta-analysis by Fraser et al (24) estimated average sensitivity and specificity of RADTs as 0.884 and 0.91 respectively. The European Society for Clinical Microbiology and Infectious Diseases’ guidelines state that throat culture is not required following negative RADT result and that throat cultures are not required for routine GAS pharyngitis diagnosis(15), but guidelines from the Infectious Diseases Society of America recommend backup throat cultures for negative RADT results in children and adolescents but not for adults(16). The latter guidelines also state that backup throat cultures are not needed for positive RADT results in children and adolescents because RADTs are highly specific.

Clinicians need to know if current practice of using Centor or McIsaac score thresholds to administer RADTs and treat RADT positive patients is good at targeting antibiotics to patients who need them and also minimizes unnecessary antibiotic prescription.

A recent review and meta-analysis compared the accuracy of Centor and McIsaac scores in primary care populations where GAS prevalence ranged between 4% and 44% (17). They concluded that a score of ≤0 for both Centor and McIsaac scores may be sufficient to rule out GAS infection but that at higher scores, a RADT is still required to rule in GAS.

This review aims to estimate the diagnostic accuracy of Centor and McIsaac scores to identify GAS pharyngitis among patients presenting to secondary care with pharyngitis. We also aim to compare expected antibiotic use under different testing scenarios, involving use of Centor or McIsaac at each threshold as a triage to RADT testing, as a tool for considering which threshold might be most appropriate.

## Methods

This study followed systematic review methods recommended by Cochrane and the Centre for Reviews and Dissemination (18, 19). A review protocol was published on PROSPERO, registration number CRD42021267413. PRIMSA-DTA guidelines were followed for reporting (20).

### Study eligibility criteria

Cohort, case control studies, and nested case control studies that included study populations consisting of adults or children presenting to secondary care with pharyngitis, that used McIsaac or Centor scores to evaluate the risk of GAS pharyngitis and used throat culture and/or rapid antigen tests as reference standards were eligible for inclusion in our review. We considered RADTs sufficiently accurate for detecting GAS pharyngitis and so included studies that used RADT as the sole reference standard. All studies had to report sufficient data for McIsaac or Centor at scores 1, 2, 3, and 4 to make a crosstabulation of people with and without GAS against each score.

### Data sources and study selection

MEDLINE, Embase and Web of Science were searched from inception until September 2022. An update search was performed in September 2022. Search terms for clinical prediction rules (CPR), including Centor and McIsaac, were combined with terms for pharyngitis and GAS (See Supplementary data for a detailed search strategy).

Two reviewers (AK and CE) independently screened titles and abstracts for relevance, followed by the detailed assessment of relevant full-text articles against the eligibility criteria, using COVIDENCE software (21). Any discrepancies were solved by discussion until a consensus was reached or by referral to a third reviewer (ME).

### Data extraction

Data extraction forms were developed in COVIDENCE (21), tested on a small sample of studies and amended where necessary. We extracted information on study type, care setting, participant recruitment, participant age, total study population, type of reference standard, McIsaac or Centor scores (index tests) at all reported thresholds, and reference standard results. Non-GAS culture results were recorded as negative GAS cultures. Studies which omitted reporting number of cases with McIsaac score = 0 and McIsaac score = 5 were assumed to have no participants with these scores. If studies combined thresholds 0 and 1 and 4 and 5, we would consider these thresholds as 1 and 4, respectively, to allow comparison between Centor and McIsaac score for thresholds 1-4. If studies reported data for two reference standards separately i.e. RADT and throat culture, we extracted and used the throat culture data. Data extraction was performed by one reviewer (AK or CE) and checked by the other. Discrepancies were resolved through discussion.

### Assessment of risk of bias

The Quality Assessment of Diagnostic Accuracy Studies (QUADAS-2) tool was used to assess the included studies’ quality which assesses 4 domains: patient selection, index test, reference standard and flow and timing (22). Low risk of bias was judged for: patient selection if participants were recruited consecutively, index test if it was interpreted without knowledge of reference standard results, and flow and timing if no data were missing and the clinical measures included in the prediction rule were measured before sample acquisition for RADT and/or throat culture. The reference standard domain was judged to be low risk in studies that used throat cultures as the reference standard and high risk if they used RADT as the only reference standard, given the limited accuracy of RADTs (23). We also judged this domain to be high risk if the reference standard was RADT followed by a confirmatory throat culture only in the RADT negatives as a proportion of patients would have only received an RADT reference standard. High risk of bias was determined for failure to meet these domain specific criteria. Domains were judged at unclear risk of bias if there was insufficient information for assessment. Studies which use convenience recruitment, inclusion/exclusion criteria specifically targeted at criteria from McIsaac or Centor score and not general pharyngitis symptoms were also judged to have applicability concerns due to potentially not recruiting a representative population of pharyngitis patients presenting to secondary care. Overall study level risk of bias was deemed low if all domains were at low risk, unclear if any domain had unclear risk of bias and no domains were at high risk, and high if any domain had high risk of bias. We presented risk of bias assessments in traffic light plots using the robvis R package (24).

### Data synthesis and analysis

We first plotted study-level receiver operator characteristic (ROC) curves for each score. MetaDTA: Diagnostic Test Accuracy Meta-Analysis v2.0 (MetaDTA), a web application (25, 26), was then used to perform meta-analyses to pool sensitivity and specificity across studies at each score threshold. MetaDTA fits binomial bivariate random effects models (27, 28), which are generalized linear mixed effect models, using the glmer function from the lme4 R package (29). MetaDTA was used to estimate pooled sensitivity, specificity, positive likelihood ratio and negative likelihood ratio for each threshold. A separate meta-analysis was performed for each score at each threshold, with summary estimates of sensitivity and specificity plotted in ROC space, together with 95% confidence ellipses and 95% prediction ellipses. Summary ROC curves (showing the pooled results across thresholds) were also added to the plots showing study-level ROC curves. Positive predictive values (PPVs) and negative predictive values (NPVs) for each score threshold were calculated using a GAS prevalence of 24.1% in secondary care(6). We also performed sensitivity analyses, restricting the meta-analyses to studies that used throat culture as the reference standard.

### Impact on antibiotic prescription

To aid interpretation of results, we considered a hypothetical cohort of 10,000 patients presenting at secondary care with pharyngitis. We assumed a prevalence of GAS of 24.1%, as has been reported for patients presenting to secondary care settings (6). We compared hypothetical scenarios in which a CPR (McIsaac or Centor score) is used as an initial triage test for RADT testing, to a baseline scenario in which all patients are given a RADT on presentation. In all scenarios, we assumed only individuals who received a positive RADT test were given antibiotics. In scenarios involving CPRs, we assumed that RADT tests were only performed if the initial CPR score was greater than or equal to each of thresholds 1, 2, 3 or 4. We assumed sensitivity of 88.4% and specificity of 91% for RADT (23) and based the sensitivity and specificity of the CPRs at each score on the results from meta-analyses restricted to studies using throat culture as the reference standard. RADT and CPR accuracy were assumed to be conditionally independent. We compared across scenarios (1) the number of GAS patients given antibiotics and (2) the number of non-GAS patients given antibiotics.

## Results

Database searches identified 784 unique records. Title and abstract screening identified 163 potentially relevant records of which 15 were included in the review (Figure 1). Of 15 included studies, eight evaluated McIsaac score (30-37) and seven Centor score (38-43). No studies evaluated both scores.

**Figure 1:**
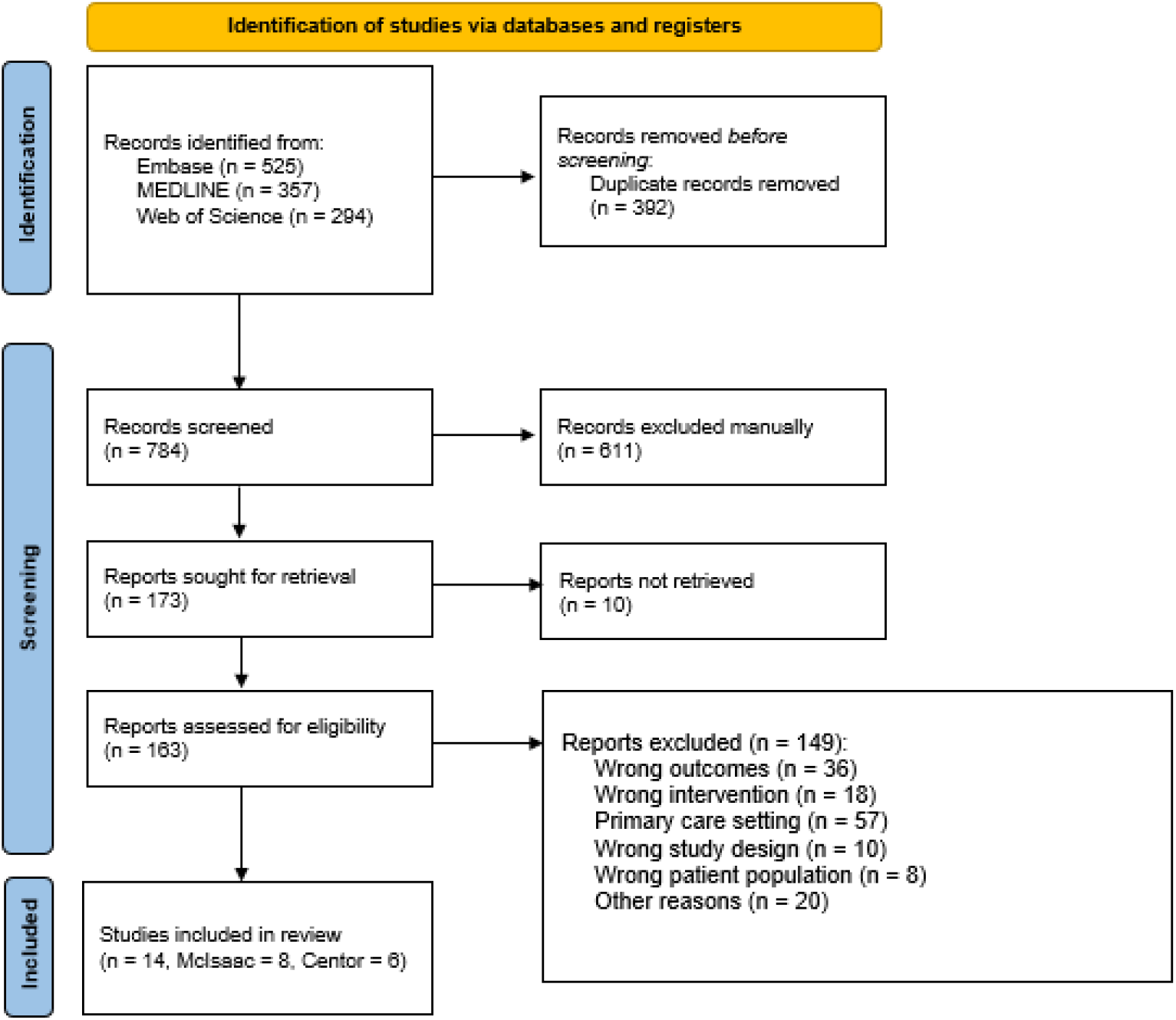
PRISMA Flow diagram of the study selection process.

### Study characteristics

Six studies have been conducted in the USA, two in Japan, and the rest in Egypt, Greece, Australia, Brazil, Croatia, Latvia, France and Pakistan (Table 1). Ten studies used both RADTs and throat cultures (30-35, 38, 39, 42, 43) as the reference standard: throat culture performed depending on RADT results (32, 35, 38, 43), or both tests performed (30, 31, 33, 34, 39, 42). Two studies (36, 41) only used RADT, and two studies (37, 40) only used throat cultures as the reference standard. Across Centor studies there were 7400 patients in total, of which Rich et al(43) and Rimoin and colleagues (44) contributed 3963 and 2472 patients respectively. A total of 4281 patients were included for the McIsaac score, with similar sample sizes across these eight studies.

**Table 1.**
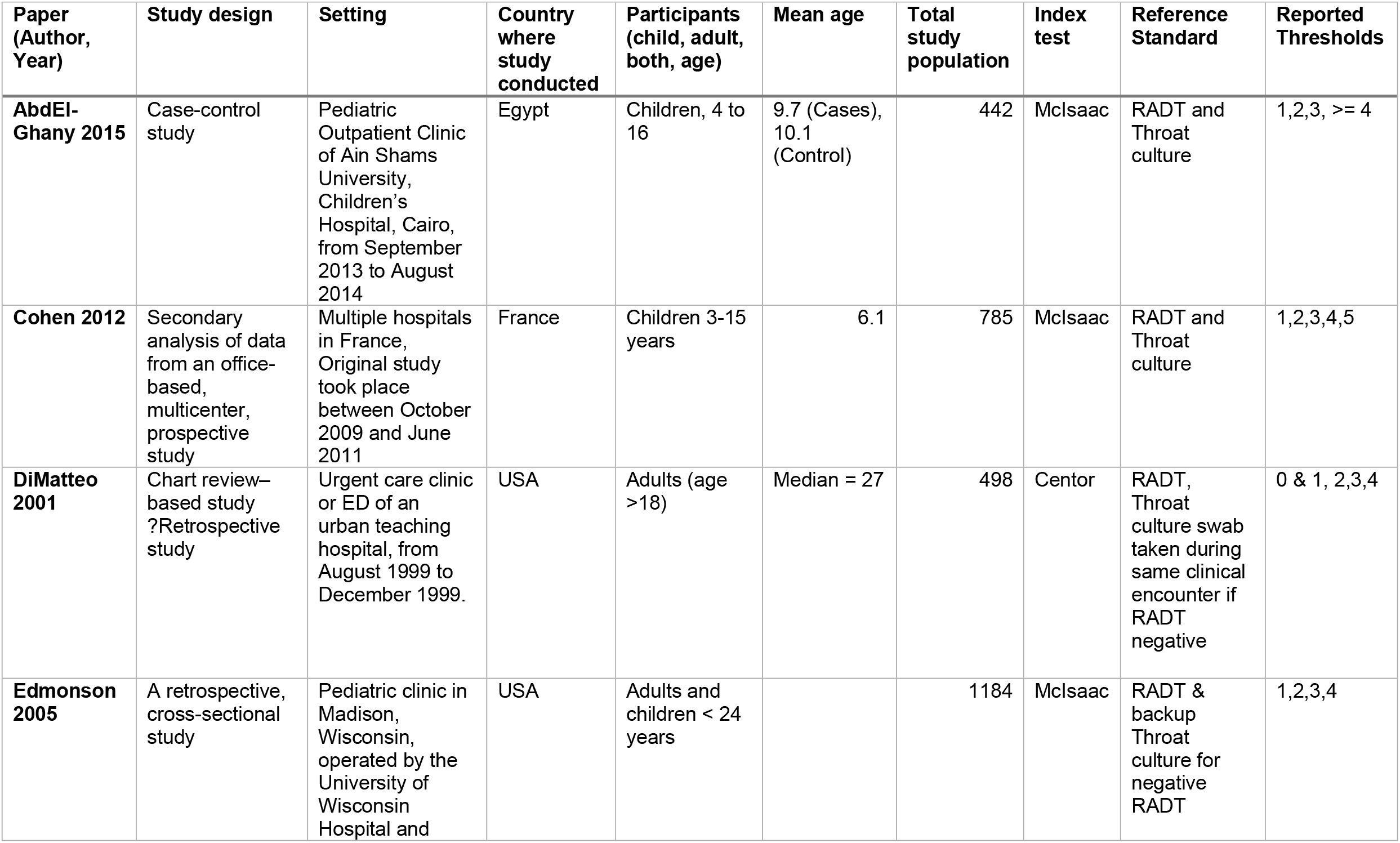
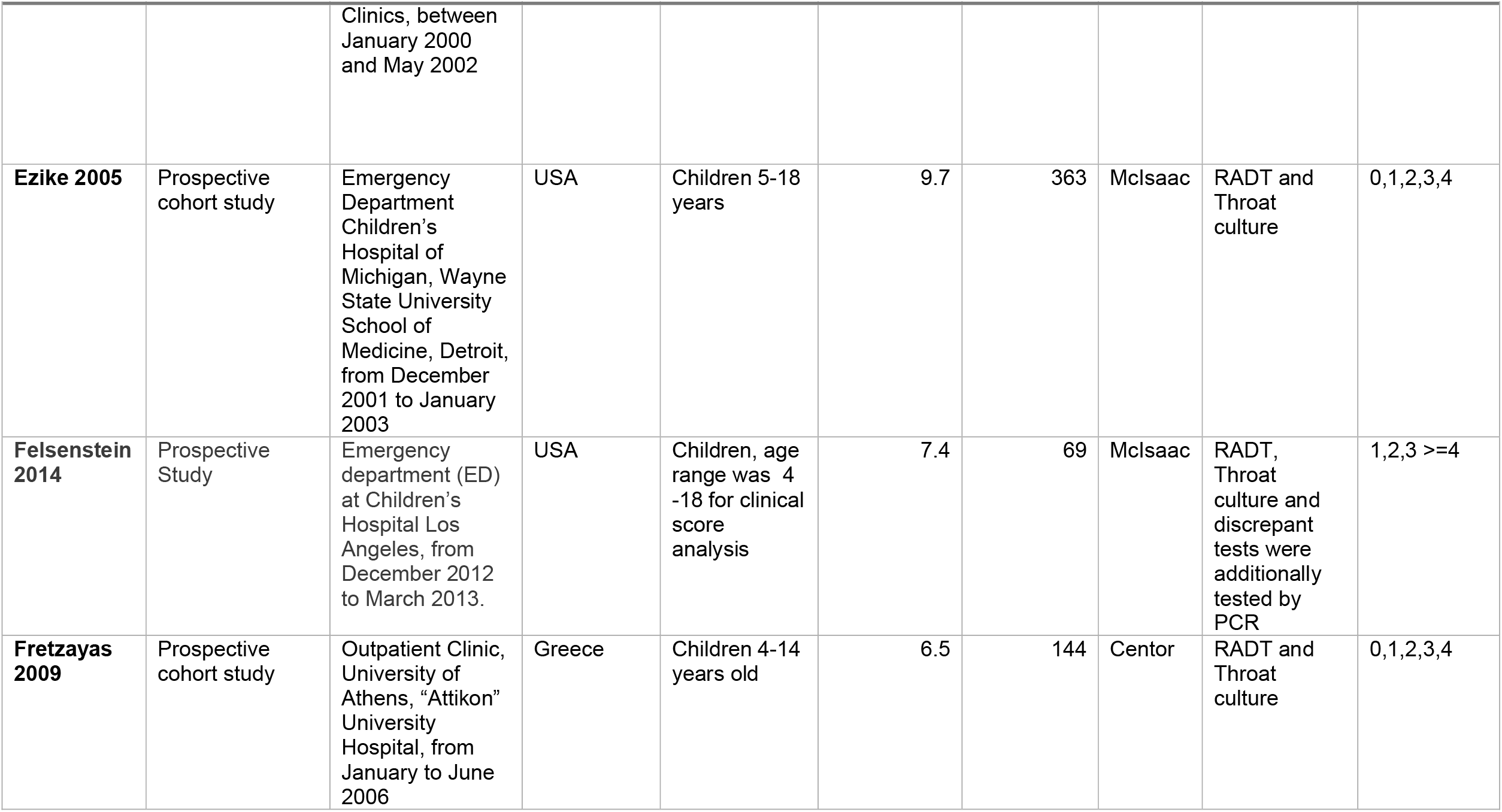
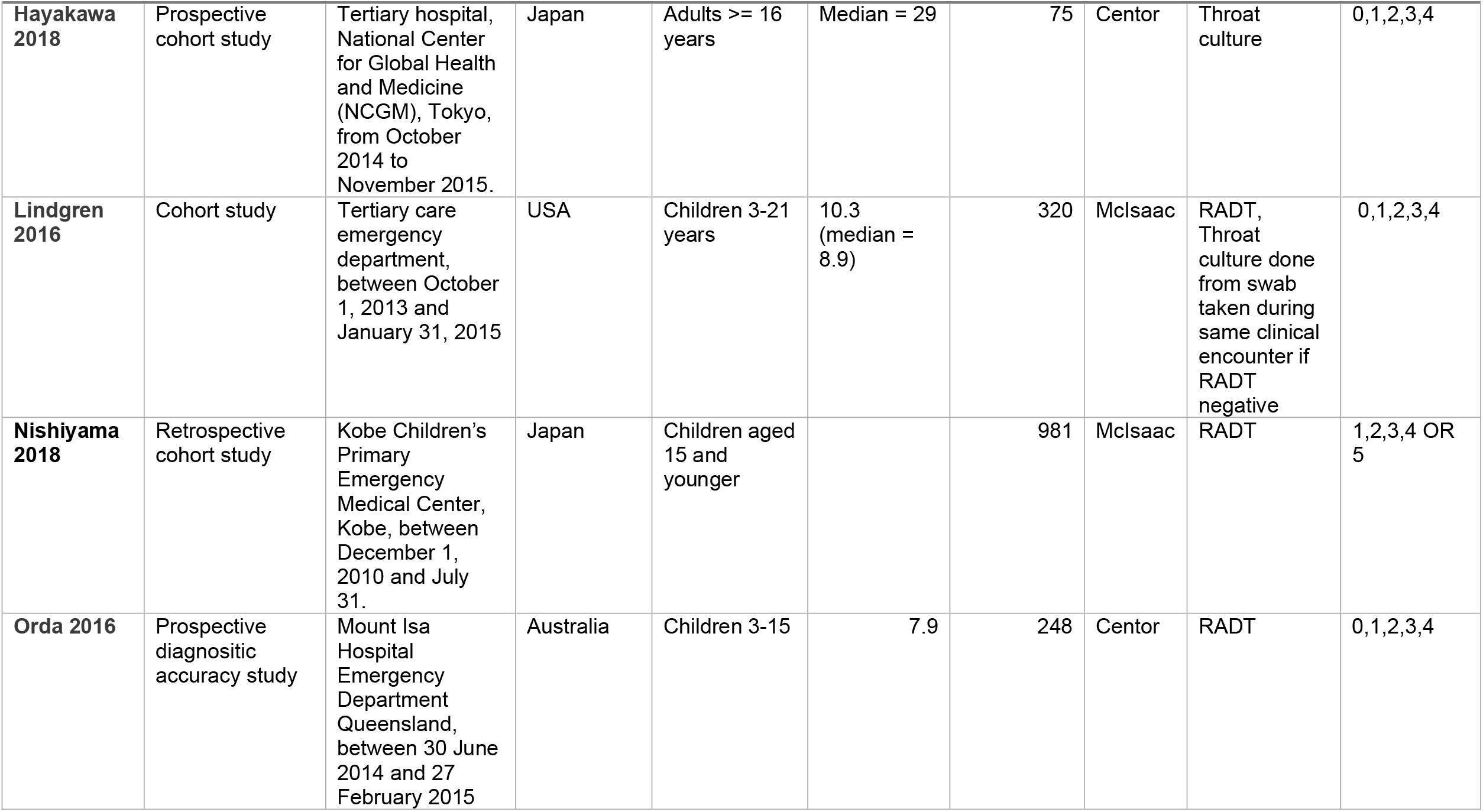
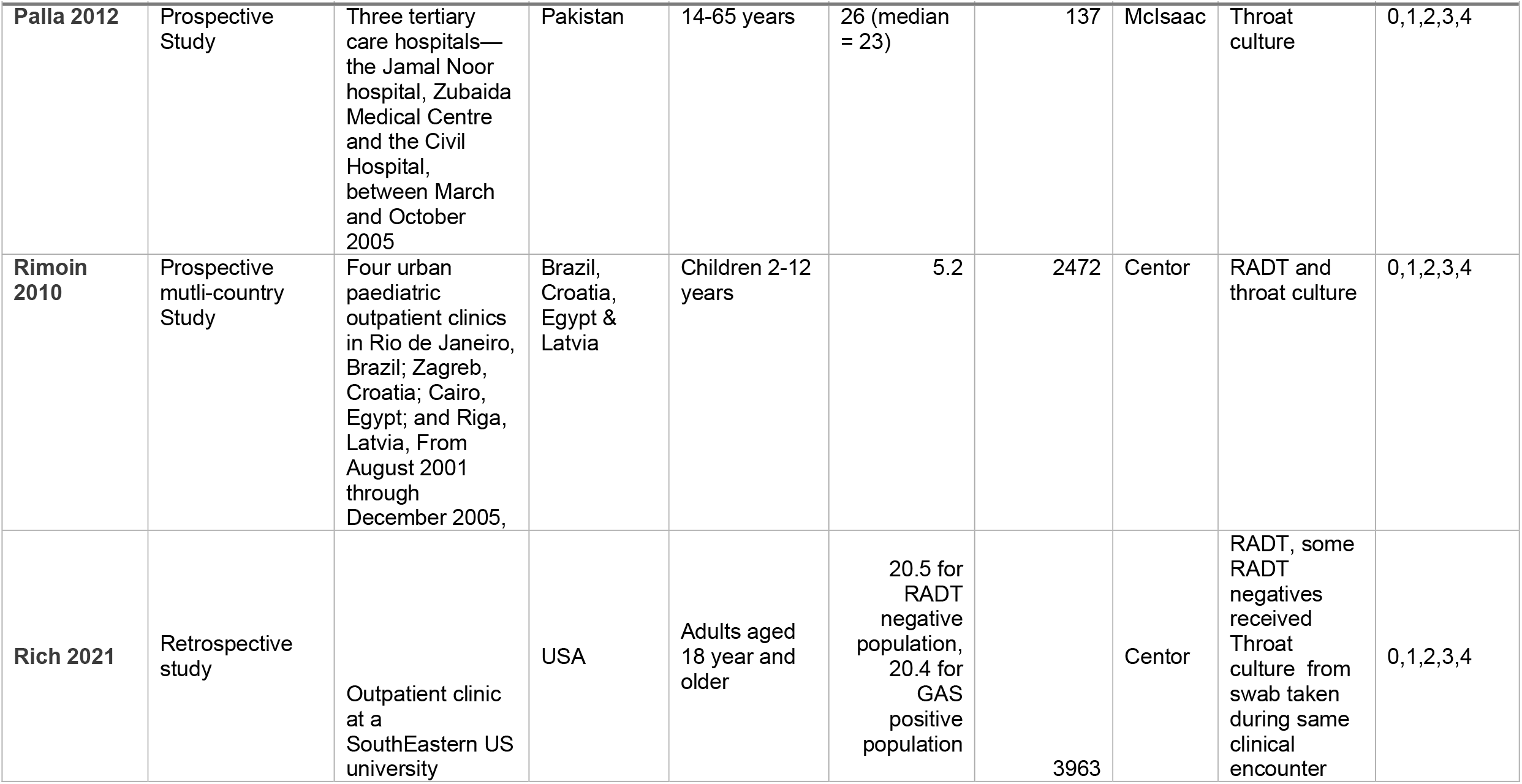
Study characteristics.

| **Paper**<br>**(Author,**<br>**Year)** | **Study design** | **Setting** | **Country**<br>**where**<br>**study**<br>**conducted** | **Participants**<br>**(child, adult,**<br>**both, age)** | **Mean age** | **Total**<br>**study**<br>**population** | **Index**<br>**test** | **Reference**<br>**Standard** | **Reported**<br>**Thresholds** |
| --- | --- | --- | --- | --- | --- | --- | --- | --- | --- |
| **AbdEl-Ghany 2015** | Case-control study | Pediatric Outpatient Clinic of Ain Shams University, Children's Hospital, Cairo, from September 2013 to August 2014 | Egypt | Children, 4 to 16 | 9.7 (Cases), 10.1 (Control) | 442 | Mclsaac | RADT and Throat culture | 1,2,3, >= 4 |
| **Cohen 2012** | Secondary analysis of data from an office-based, multicenter, prospective study | Multiple hospitals in France, Original study took place between October 2009 and June 2011 | France | Children 3-15 years | 6.1 | 785 | Mclsaac | RADT and Throat culture | 1,2,3,4,5 |
| **DiMatteo 2001** | Chart review–based study<br>?Retrospective study | Urgent care clinic or ED of an urban teaching hospital, from August 1999 to December 1999. | USA | Adults (age >18) | Median = 27 | 498 | Centor | RADT, Throat culture swab taken during same clinical encounter if RADT negative | 0 & 1, 2,3,4 |
| **Edmonson 2005** | A retrospective, cross-sectional study | Pediatric clinic in Madison, Wisconsin, operated by the University of Wisconsin Hospital and | USA | Adults and children < 24 years |  | 1184 | Mclsaac | RADT & backup Throat culture for negative RADT | 1,2,3,4 |

| Paper (Author, Year) | Study design | Setting | Country where study conducted | Participants (child, adult, both, age) | Mean age | Total study population | Index test | Reference Standard | Reported Thresholds |
| --- | --- | --- | --- | --- | --- | --- | --- | --- | --- |
|  |  | Clinics, between January 2000 and May 2002 |  |  |  |  |  |  |  |
| **Ezike 2005** | Prospective cohort study | Emergency Department Children's Hospital of Michigan, Wayne State University School of Medicine, Detroit, from December 2001 to January 2003 | USA | Children 5-18 years | 9.7 | 363 | Mclsaac | RADT and Throat culture | 0,1,2,3,4 |
| **Felsenstein 2014** | Prospective Study | Emergency department (ED) at Children's Hospital Los Angeles, from December 2012 to March 2013. | USA | Children, age range was 4 -18 for clinical score analysis | 7.4 | 69 | Mclsaac | RADT, Throat culture and discrepant tests were additionally tested by PCR | 1,2,3 >=4 |
| **Fretzayas 2009** | Prospective cohort study | Outpatient Clinic, University of Athens, "Attikon" University Hospital, from January to June 2006 | Greece | Children 4-14 years old | 6.5 | 144 | Centor | RADT and Throat culture | 0,1,2,3,4 |
| **Hayakawa**<br>**2018** | Prospective<br>cohort study | Tertiary hospital,<br>National Center<br>for Global Health<br>and Medicine<br>(NCGM), Tokyo,<br>from October<br>2014 to<br>November 2015. | Japan | Adults >= 16<br>years | Median = 29 | 75 | Centor | Throat<br>culture | 0,1,2,3,4 |
| **Lindgren**<br>**2016** | Cohort study | Tertiary care<br>emergency<br>department,<br>between October<br>1, 2013 and<br>January 31, 2015 | USA | Children 3-21<br>years | 10.3<br>(median =<br>8.9) | 320 | Mclsaac | RADT,<br>Throat<br>culture done<br>from swab<br>taken during<br>same clinical<br>encounter if<br>RADT<br>negative | 0,1,2,3,4 |
| **Nishiyama**<br>**2018** | Retrospective<br>cohort study | Kobe Children's<br>Primary<br>Emergency<br>Medical Center,<br>Kobe, between<br>December 1,<br>2010 and July<br>31. | Japan | Children aged<br>15 and<br>younger |  | 981 | Mclsaac | RADT | 1,2,3,4 OR<br>5 |
| **Orda 2016** | Prospective<br>diagnostic<br>accuracy study | Mount Isa<br>Hospital<br>Emergency<br>Department<br>Queensland,<br>between 30 June<br>2014 and 27<br>February 2015 | Australia | Children 3-15 | 7.9 | 248 | Centor | RADT | 0,1,2,3,4 |
| **Palla 2012** | Prospective<br>Study | Three tertiary<br>care hospitals—<br>the Jamal Noor<br>hospital, Zubaida<br>Medical Centre<br>and the Civil<br>Hospital,<br>between March<br>and October<br>2005 | Pakistan | 14-65 years | 26 (median<br>= 23) | 137 | Mclsaac | Throat<br>culture | 0,1,2,3,4 |
| **Rimoin**<br>**2010** | Prospective<br>mutli-country<br>Study | Four urban<br>paediatric<br>outpatient clinics<br>in Rio de Janeiro,<br>Brazil; Zagreb,<br>Croatia; Cairo,<br>Egypt; and Riga,<br>Latvia, From<br>August 2001<br>through<br>December 2005, | Brazil,<br>Croatia,<br>Egypt &<br>Latvia | Children 2-12<br>years | 5.2 | 2472 | Centor | RADT and<br>throat culture | 0,1,2,3,4 |
| **Rich 2021** | Retrospective<br>study | Outpatient clinic<br>at a<br>SouthEastern US<br>university | USA | Adults aged<br>18 year and<br>older | 20.5 for<br>RADT<br>negative<br>population,<br>20.4 for<br>GAS<br>positive<br>population | 3963 | Centor | RADT, some<br>RADT<br>negatives<br>received<br>Throat<br>culture from<br>swab taken<br>during same<br>clinical<br>encounter | 0,1,2,3,4 |

Studies included both adults and children and also used different age ranges to classify children and adults – e.g. Hayakawa et al (40) defined adults as those aged 16 or above, while El-Ghany et al (30) defined adults as being over 18 years old.

### Risk of bias assessment

Only two studies(44) explicitly mentioned the use of consecutive patient recruitment and were therefore assessed as low risk of bias for the patient selection domain. Seven out of eight McIsaac studies (Figure 2a) and three out of seven Centor studies (Figure 2b) were judged to be at unclear risk of bias overall due to patient recruitment method not being described. The study by DiMatteo and colleagues(38) was also judged at unclear risk of bias for using consecutively recruiting retrospective patients who received a RADT, as it was not described if all patients presenting to the hospital with pharyngitis received a RADT.

**Figure 2a,b:**
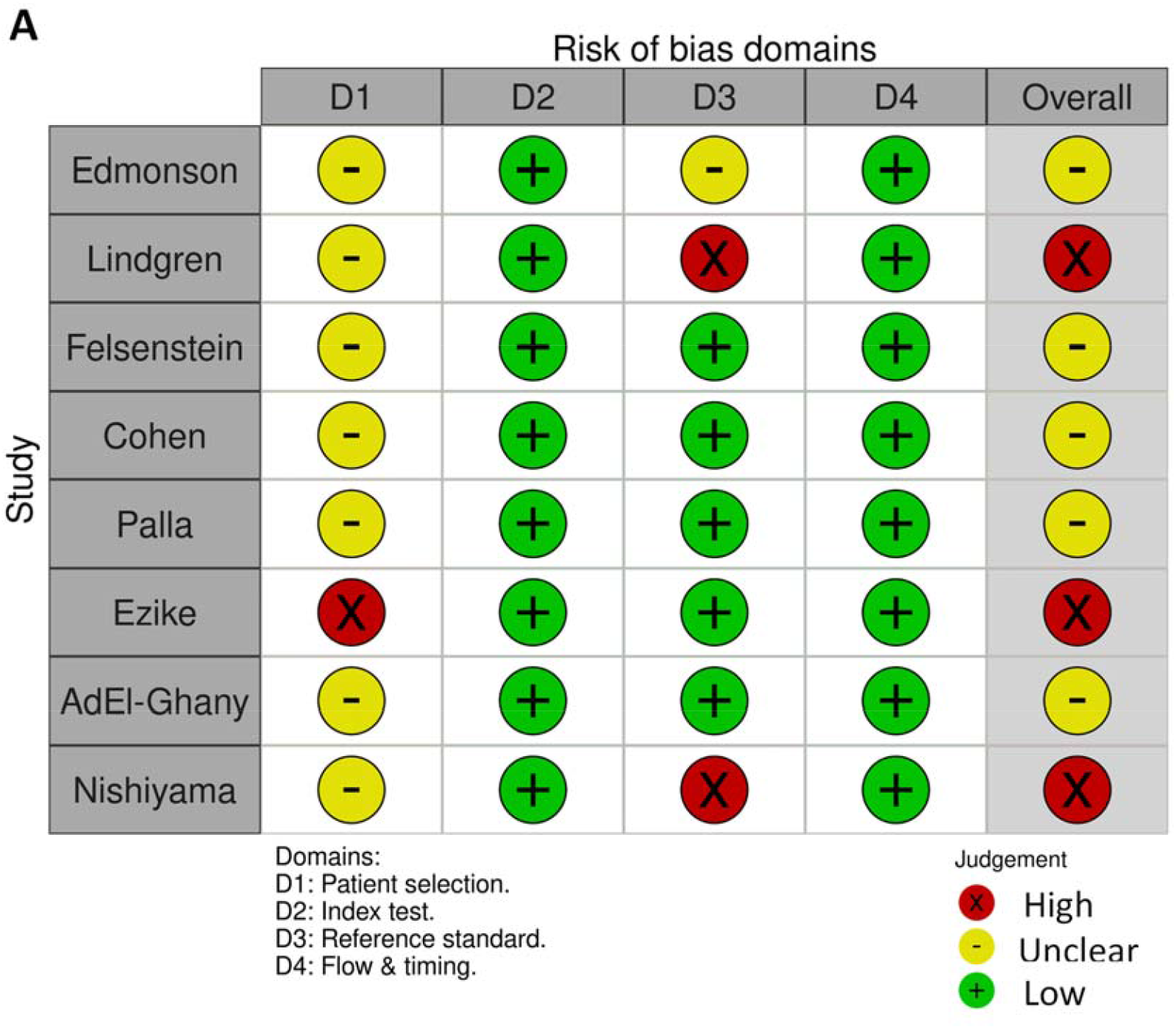
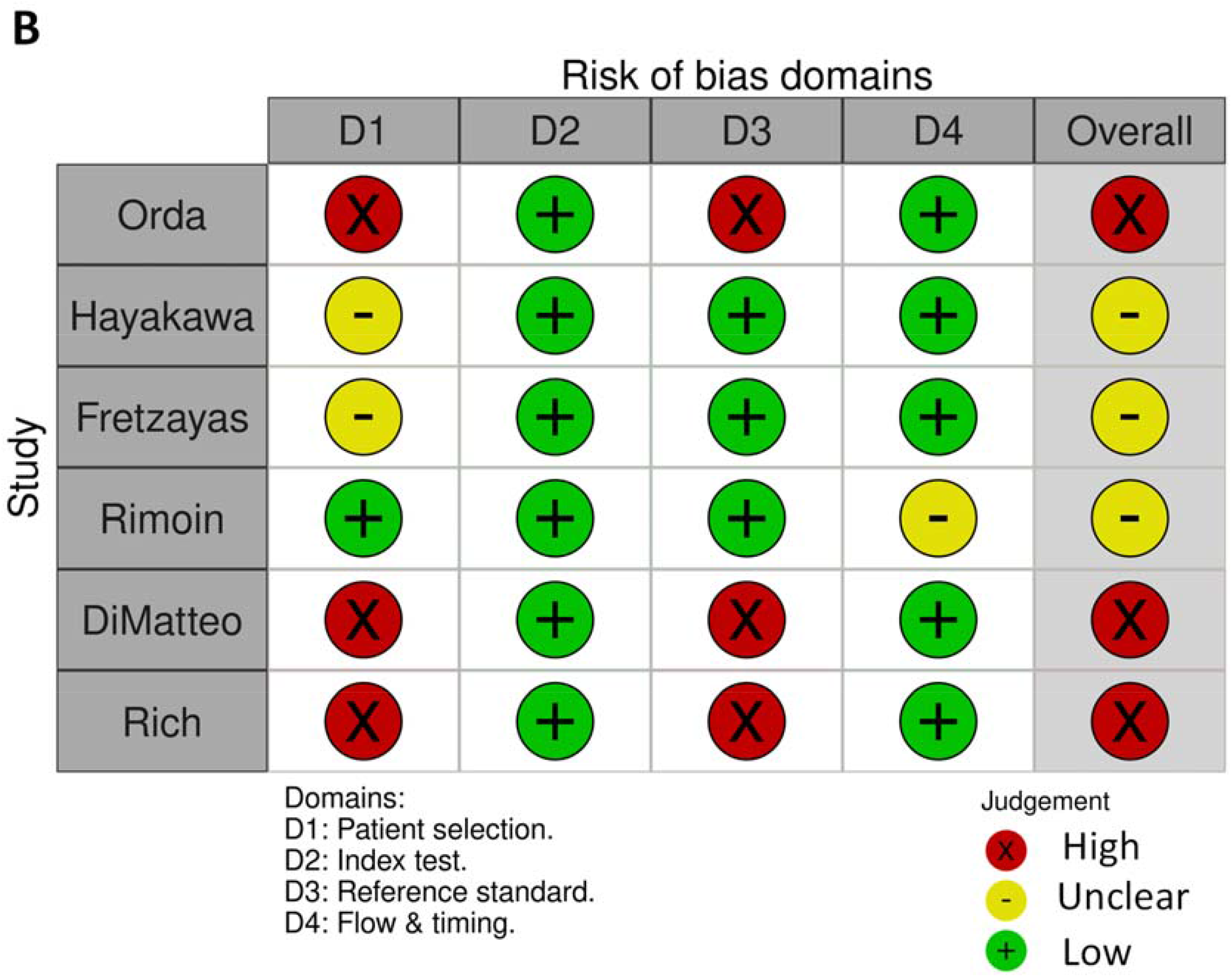
**Risk of bias traffic light plots of the QUADAS-2 risk of bias assessment results for McIsaac studies (1a) and Centor studies (1b) produced using the robVis web application**

Eight studies used at least two reference standards. These studies were considered low risk of bias for flow and timing because all patients had throat swabs taken at the same clinical encounter for both RADT and backup throat culture. Patient exclusion from analysis was only done for appropriate reasons, such as missing clinical or reference standard data, so flow and timing domain was mostly low risk. One study(42) was an exception and was of unclear risk of bias, because 4.8% of data were not included in the analysis due to lost laboratory data. We could not establish the exact nature of the patient data that was lost (e.g. whether all were from patients from a particular area/ socioeconomic background, etc.) and so we could not determine how this affected the risk of bias for the flow and timing domain.

### Sensitivity and specificity of McIsaac and Centor scores

Sensitivity and specificity per study per threshold are shown for Centor score in Figure 3A and McIsaac score in Figure 3B. For McIsaac, Ezike et al also has two points lying just below the AUROC = 0.5 line, while Felsenstein et al has two points lying on this line

**Figures 3a,b:**
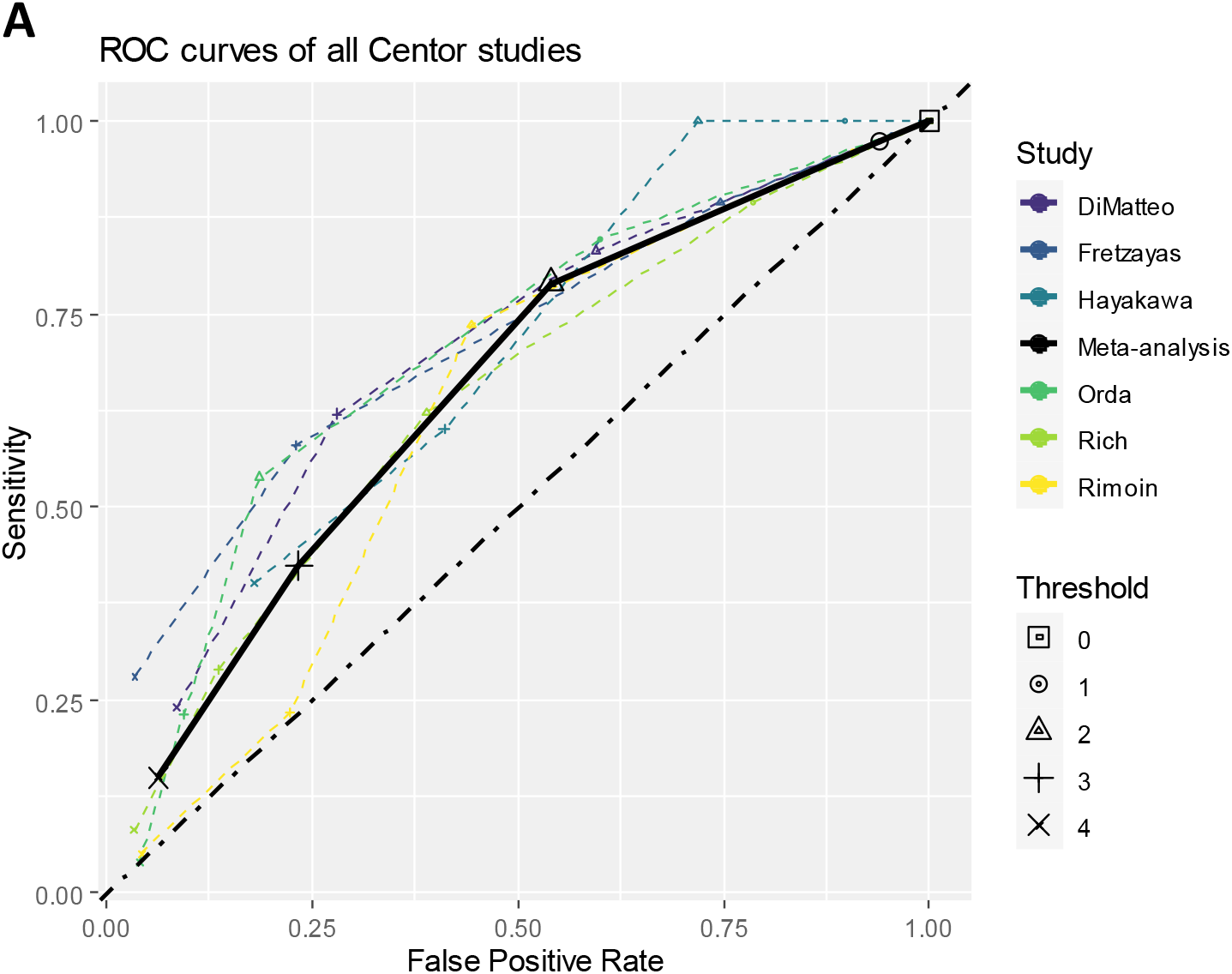
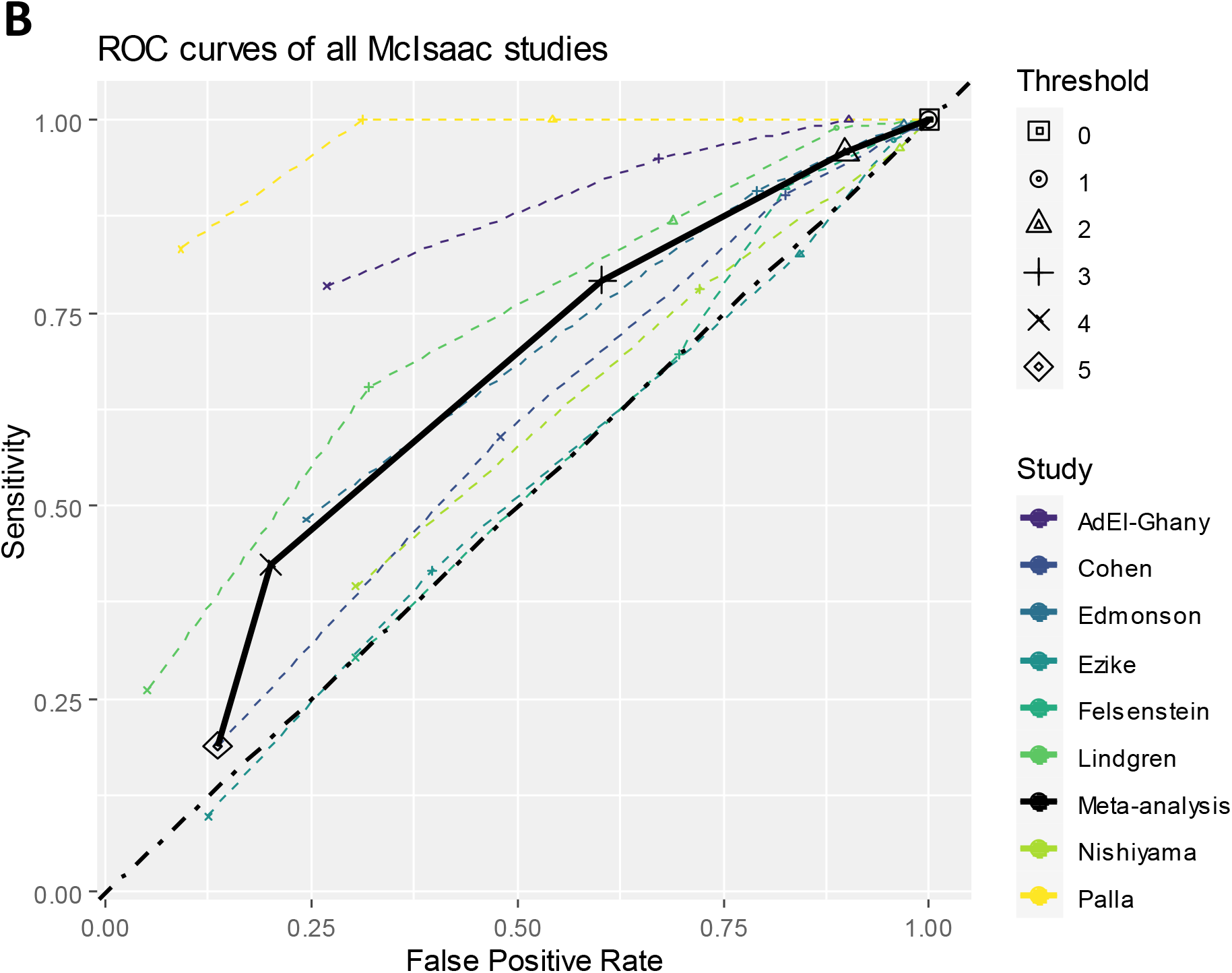
Sensitivity versus 1-specificity (false positive rate) estimates per threshold per study plotted in receiver operating characteristic (ROC) space of the studies for Centor (3a) and McIsaac (3b) studies for all thresholds. Studies are colour coded and different symbols are used to indicate different numerical thresholds. The solid black line represents the summary estimate from the meta-analysis. The black dot-dash line represents the gradient of a clinical prediction rule with 50% sensitivity and 50% specificity.

Centor and McIsaac score pooled sensitivity, specificity, positive predictive value (PPV) and negative predictive value (NPV) for a GAS prevalence of 24.1%, positive likelihood ratio (+LR) and negative likelihood ratio (-LR) are shown in Table 2a. Supplementary Figure 1A-D show results from each of the eight meta-analyses, including confidence and prediction ellipses. McIsaac score has higher summary sensitivity and lower summary specificity than the same Centor score thresholds (Table 2a). Visual comparison shows that the 95% confidence and prediction regions overlap across both scores (except for the 95% confidence region for threshold ≥3). The wide confidence intervals and correspondingly large confidence regions at most thresholds indicate high uncertainty in the summary estimates, and the much larger prediction regions indicate substantial heterogeneity across studies, for each threshold for both scores.

**Table 2A).**
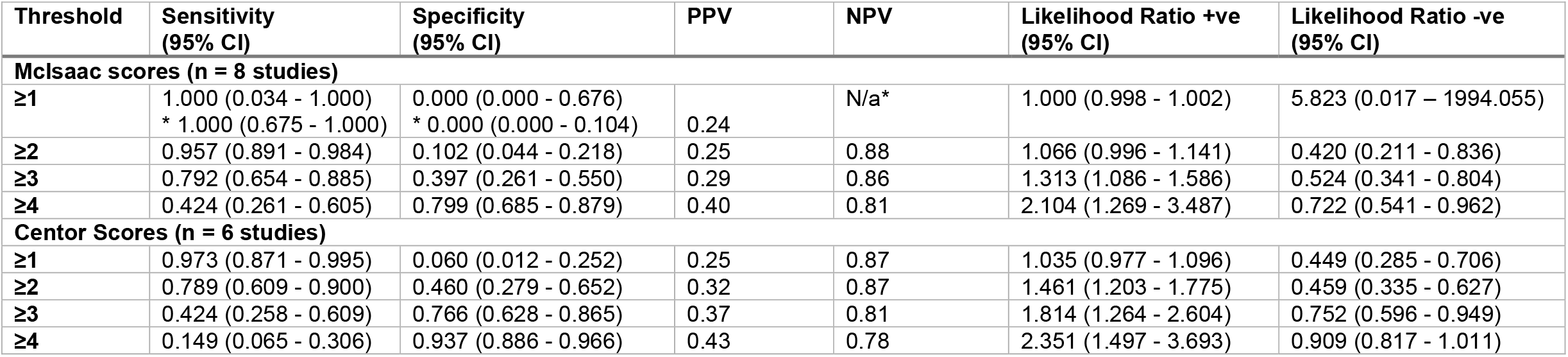
Sensitivity and specificity of McIsaac and Centor scores. Summary estimates of sensitivity and specificity, positive and negative likelihood ratios for McIsaac and Centor scores at several thresholds. PPV and NPV calculated assuming a GAS prevalence of 24.1% in secondary care(6). CI = confidence interval. *Results from sensitivity analysis using metandi in Stata, after noting unintuitively wide 95% CIs from the primary analysis. * NPV could not be calculated due to true negative and false negative cases being zero Abbreviations: PPV = Positive Predictive Value, NPV = Negative Predictive Value

**B).** Comparison of positive predictive values (PPV) for McIsaac and Centor scores per threshold from this review and existing literature. PPV for Willis et al (systematic review in primary care) was calculated from data presented in the paper. This review’s mean Centor study GAS prevalence = 27%, mean McIsaac study GAS prevalence = 37%, Original Centor study(10) GAS prevalence = 17%, Original McIsaac study(11) GAS prevalence = 13.8%, Willis et al(17) systematic review median GAS prevalence for Centor score = 26.4%, for McIsaac score = 23%, Fischer Walker et al(45) GAS prevalence = 24.6%.

| Threshold | This review's PPV (%) | Original Study PPV(10) (11)(%) | Fischer Walker et al (45)PPV (%) | Willis et al (17)PPV (%) |
| --- | --- | --- | --- | --- |
| Mclsaac scores |  |  |  |  |
| 0 | 24 | 2.5 | - | 28 |
| 1 | 24 | 5.1 | - | 25 |
| 2 | 25 | 11.2 | 28 | 28 |
| 3 | 29 | 27.8 | 37 | 34 |
| 4 | 40 | 52.8 | 40 | 44 |
| 5 | 31 | N/A | - | 57 |
| Centor scores |  |  |  |  |
| 0 | 24 | 2.5 | - | - |
| 1 | 25 | 6.5 | 25 | 28 |
| 2 | 32 | 15 | 33 | 32 |
| 3 | 37 | 32 | 44 | 41 |
| 4 | 43 | 56 | - | 55 |

Results from sensitivity analyses restricting to studies that used throat culture as the reference standard are shown in Supplementary Table 1. Results were comparable between both analyses, with overlapping 95% Cis.

### Impact on antibiotic prescription

The estimations of how many antibiotic prescriptions could be prevented with the use of McIsaac or Centor scores as a triage to RADT, compared with using RADT alone, to prescribe antibiotics and how many patients would not receive antibiotics who may need them are presented in Table 3, for McIsaac and Centor scores at each threshold.

**Table 3.** Impact of two-stage clinical assessment strategy using McIsaac and Centor score thresholds and RADT on antibiotic prescription in a hypothetical population. Estimated numbers of antibiotic prescriptions among a hypothetical population of 10,000 patients presenting to secondary care with pharyngitis, including 2,410 (i.e. 24.1% prevalence) patients with presence of GAS, using the pooled estimates of sensitivity and specificities per McIsaac and Centor score threshold calculated from studies which used throat culture as the reference standard (Supplementary table 1), and assuming sensitivity and specificity of 88.4% and 91% respectively for RADT. Abbreviations: GAS = Group A Streptococcus, RADT = Rapid Antigen Diagnostic Test.

| Threshold | Sensitivity | Specificity | Step 1: classification based on score |  | Step 2: RADT applied after positive score threshold |  |  |  |
| --- | --- | --- | --- | --- | --- | --- | --- | --- |
|  |  |  | Number of GAS cases correctly identified by threshold score (1) | Number of non-GAS cases that are FP using threshold(2) | Number of GAS cases who receive antibiotics (3) | Number of non-GAS patients that receive antibiotics (4) | Reduction in number of GAS cases receiving antibiotics, compared with baseline scenario (5)* | Reduction in number of non-GAS cases receiving antibiotics, compared with baseline scenario (6) * |
| Baseline scenario (RADT for all) * |  |  | N/A | N/A | 2130 | 683 |  |  |
| Mclsaac scores |  |  |  |  |  |  |  |  |
| ≥1 | 0.998 | 0.001 | 2405 | 7579 | 2126 | 682 | **4** | **1** |
| ≥2 | 0.959 | 0.125 | 2311 | 6641 | 2043 | 598 | **87** | **85** |
| ≥3 | 0.819 | 0.410 | 1974 | 4478 | 1745 | 403 | **386** | **280** |
| ≥4 | 0.478 | 0.776 | 1152 | 1700 | 1018 | 153 | **1112** | **530** |
| Centor scores |  |  |  |  |  |  |  |  |
| ≥1 | 0.984 | 0.052 | 2371 | 7195 | 2096 | 648 | **34** | **36** |
| ≥2 | 0.903 | 0.237 | 2176 | 5791 | 1924 | 521 | **207** | **162** |
| ≥3 | 0.581 | 0.516 | 1400 | 3674 | 1238 | 331 | **893** | **352** |
| 4 | 0.290 | 0.853 | 699 | 1116 | 618 | 100 | **1513** | **583** |

- (1) Number of GAS patients 'positive' at this threshold = $10,000 * \text{Prevalence} * \text{Sensitivity}$ - (2) Number of non-GAS patients 'positive' at this threshold = $10,000 * (1 - \text{Prevalence}) * (1 - \text{Specificity})$ - (3) Number of GAS patients who receive antibiotics after RADT = (column 1)\*(RADT sensitivity) - (4) Number of non-GAS patients who receive antibiotics after RADT = (column 2)\*(1 – RADT specificity) - (5) Reduction in number of GAS patients receiving antibiotics due to the threshold step (relative to strategy of RADT test for everyone) = $(10,000 * \text{Prevalence} * \text{RADT sensitivity}) - (\text{column 3})$ - (6) Reduction in number of non-GAS patients receiving antibiotics due to the threshold step (relative to strategy of RADT test for everyone) = $(10,000 * (1 - \text{Prevalence}) * (1 - \text{RADT specificity})) - (\text{column 4})$

The estimated number of false positive cases identified by the threshold step and the number of these who would be given antibiotics if a positive RADT is required is also shown, to appreciate the potential impact of using additional RADTs on unnecessary prescription.

Using CPRs as an initial triage step reduces the number of antibiotic prescriptions, but also reduces the number of GAS cases – who can benefit from the antibiotics – not receiving them. For example, in the baseline strategy of a RADT for everyone, 88% of GAS patients and 9% of non-GAS patients receive antibiotics. Following NICE treatment guidelines (Centor ≥3 as an initial triage for RADT testing), we estimate that only 51% of GAS patients and 4% of non-GAS patients would receive antibiotics. In our hypothetical cohort of 10,000 patients with 24.1% GAS prevalence, this translates to a reduction in number of antibiotic prescriptions from 2813 to 1569 (reduction of 1244), but we estimate that 893 of these people not prescribed would in fact have GAS pharyngitis. Similarly for the Danish (McIsaac ≥2) and German (McIsaac ≥3) treatment guidelines, we estimate that 84.8% and 72.4% of GAS pharyngitis patients and 7.9% and 5.3% of non-GAS patients would receive antibiotics, with a reduction in the number of prescriptions by 172 and 666 respectively, where 87 and 386 cases of these avoided prescriptions would have had GAS pharyngitis.

## Discussion

### Main findings

McIsaac and Centor scores aim to accurately distinguish between cases of GAS and non-GAS pharyngitis in patients presenting with pharyngitis to reduce unnecessary use of antibiotics through better targeted prescription. After reviewing studies set in secondary care investigating the ability of McIsaac and Centor scores to distinguish between GAS pharyngitis and non-GAS pharyngitis, we note that there is little separating McIsaac and Centor scores once it is appreciated that McIsaac score of 0 or 1 is equivalent to Centor score of 0, McIsaac score of 2 is equivalent to Centor score of 1 and so on (Tables 2 & 3). We find that different treatment thresholds need to be used for these scores.

We estimate that Centor scores of ≥3 and 4 have sensitivities of 58% and 29%, respectively, compared to McIsaac score ≥2 and ≥3 which have sensitivities of 96% and 82%, respectively. Current NICE guideline recommendations therefore potentially misses a large cohort of GAS positive pharyngitis patients.

Using clinical scores either alone or as a triage to RADT testing leaves a sizable proportion of GAS positive patients missing out on antibiotics that could have benefited from them. RADT use on threshold positive patients significantly limits the incorrect treatment of false positive cases, particularly at low thresholds, and thereby prevents unnecessary use of antibiotics. However, the low sensitivity of CPR scores when implemented at higher cutoffs as a triage step leads to a significant proportion of GAS cases not receiving a RADT test and therefore not receiving treatment. Therefore, RADT follow up of patients with low CPRs is likely to have the most clinical value.

### Strength and limitations

Our systematic review was carried out according to the best practice recommendations for review conduct. Our search strategy was comprehensive, study selection and data extraction were performed by one reviewer (AK or CE) and checked by the other, and the review was reported according to PRISMA-DTA guidelines.

However, we believe that there are significant limitations to the evidence base we examined in this review. Firstly, studies used heterogeneous age ranges for adults and children, preventing stratified analyses based on age groups. Secondly, several included studies solely used RADTs as the reference standard instead of more accurate throat cultures – although our sensitivity analyses restricting to studies using throat culture as the reference standard provided similar results. Thirdly, RADTs or throat cultures do not distinguish between GAS commensal carriage and GAS infection, meaning that identifying true positive GAS pharyngitis where the causative organism is GAS is difficult and the number of true positives and false positive is uncertain.

### Comparisons with existing literature

We compared our findings to those from existing literature (Table 2b). Compared to the original Centor study(10) conducted on adults presenting to an urban emergency room and the original McIsaac study(11) conducted on patients aged 3 to 76 presenting to a primary care centre, we report higher PPVs for thresholds Centor 0-3 and lower PPV for Centor = 4. For McIsaac score we report higher PPVs for thresholds 0-3, and lower PPV for threshold ≥4.

Fischer Walker and colleagues(45) compared multiple CPRs (Breese score, Wald score, Centor score, McIsaac score, Abu Reesh score, Steinhoff score, WHO score) on the same cohort of children presenting to the hospital with sore throat and pharyngeal erythema to predict streptococcal pharyngitis. Their results show lower McIsaac score sensitivity, both higher and lower Centor score sensitivity and higher specificity for both scores across equivalent thresholds, potentially due the study population being children exclusively.

Willis and colleagues evaluated Centor and McIsaac score in primary care (17); their results were comparable to this review’s findings (Table 2b): The 95% confidence intervals for McIsaac scores 0-4 and Centor scores 1-4 overlap between this review and Willis and colleagues’ review. They concluded that for both tests, “a score of ≤0 may be sufficient to rule out infection” but that higher scores could result in overuse of antibiotics and therefore justifies the implementation of additional point of care tests to confirm the diagnosis.

The Primary care Streptococcal Management (PRISM) study by Little and colleagues(46) evaluated the use of clinical scores, RADTs and delayed prescriptions in the context of pharyngitis. They found that the tested RADTs were 100% specific for GAS, but sensitivity ranged from 62% to 95%. They developed and tested the FeverPAIN score, which is based on fever, purulence, attend rapidly (within 3 days), severe inflammation and no cough or coryza. In the two primary care study cohorts included in PRISM with prevalence of 34% and 40% of A, C or G beta-haemolytic streptococci, FeverPAIN had a higher bootstrapped estimated AUROC (0.73 and 0.71) compared to the estimates from Willis and colleagues for Centor score, 0.6888 (95% CI 0.653 - 0.724), and McIsaac score, 0.7052 (95% CI 0.624 - 0.778), in primary care populations with a prevalence of 26.4% (range: 4.7%–42.0%) across Centor score studies, and 23.0% (range: 12.7%–44.8%) for McIsaac score studies (17). We did not identify any studies that have used FeverPAIN in a secondary care population. Little et al also found that symptom management and antibiotic use wise there were no reason to justify use of RADT over clinical score alone from a time and economic cost perspective and that both interventions reduced antibiotic use.

### Recommendations for practice

Risks of treating false negative and false positive cases must be considered when setting a treatment threshold: the former could lead to patients developing complications while the latter could lead to over-prescription of antibiotics and eventually increased anti-microbial resistance. The diagnostic pathway must be both highly sensitive and specific for GAS pharyngitis to treat true positive cases and minimise unnecessary antibiotic prescriptions and medicalisation of relative minor symptoms. We cannot recommend the use of one score over the other to triage patients presenting to hospitals with pharyngitis.

Adequate safety netting by clinicians would ensure that patients return should their symptoms worsen. This may allow the use of higher, less sensitive score thresholds, which exclude more GAS pharyngitis patients from receiving antibiotics at the time primary presentation while minimising potential harm to patients who miss out on receiving antibiotics, thus decreasing the number of false positives treated with antibiotics and limits medicalisation of relative minor symptoms. We cannot recommend a sufficiently inclusive threshold for both these scores to decide whether to treat patients with antibiotics or not when we regard the impact on antimicrobial stewardship with the data analysed in this review. Thus, we recommend that current antibiotic prescription guidelines are followed and clinicians safety net patients to return on worsen of/ non-resolution of symptoms.

Due to insufficient clinical effectiveness and economic evidence on the effect of using RADT after a low McIsaac or Centor on antibiotic prescriptions, we cannot make a strong recommendation for this strategy. However, our analysis is promising and so we recommend further research. Both scores have low sensitivities at high thresholds, hence we recommend that if these scores are used as a triage to RADT testing then a low threshold should be implemented for RADT testing.

### Conclusions

Centor and McIsaac scores are equally ineffective at triaging patients who need antibiotics presenting with pharyngitis at hospitals. At high thresholds too many true positive cases are missed while at low thresholds too many false positives are identified and treated leading to over-prescription of antibiotics. The former may be compensated by proper safety netting to seek help if symptoms worsen and/or delayed prescriptions by clinicians. As shown here, additional testing of positive threshold cases by RADT decreases false positives at low thresholds but at an economic cost and at the cost of further decreasing the number of true positive cases treated at high thresholds. For now, current guidelines should be followed but we recommend further study of the clinical and economic impact of RADT testing of patients with low CPR scores to determine antibiotic prescription and similar evaluation of using FeverPAIN in populations of patients presenting to hospitals. Better CPRs may be developed by combining signs and symptoms with test results, such as RADT or inflammation markers. However, the cost-effectiveness of these strategies needs to be considered carefully because they could lead to an increase in testing.

## Data Availability

All data produced in the present study are available upon reasonable request to the authors

