## Supplementary for "Systematic review and meta-analysis of the accuracy of McIsaac and Centor score in patients presenting to secondary care with Pharyngitis"

### Supplementary data

#### 1.0. Search strategy:

##### Web of Science (WoS)

SCI-EXPANDED, CPCI-S, ESCI (All years)

#1 TOPIC: (Feverpain* OR McIsaac* OR centor*) 183
#2 TOPIC: (strep* NEAR/3 (throat or throats or tonsil* or pharyn* or nasopharyn* or epipharyn* or rhinopharyn*)) 3653

#3 TOPIC: “GAS pharyngiti*” 187

#4 (#2 OR #3) 3696

#5 TOPIC: (pharyngiti* or epipharyngiti* or nasopharyngiti* or rhinopharyngiti* or pharyngalgi* or pharyngotonsilliti* or tonsilliti*) 9583

#6 TOPIC: ((throat or throats or tonsil* or pharyn* or nasopharyn* or epipharyng* or rhinopharyng*) NEAR/3 (ache or aches or aching or infect* or pain* or sore*)) 10193

#7 TOPIC: (throat near/2 (culture or cultures or swab or swabs)) 3161

#8 (#5 or #6 or #7) 20070

#9 TOPIC: (bacteria* NEAR infect* NEAR diagnos*) 38737

#10 TOPIC: (strep* NEAR/3 infect*) 11434

#11 TOPIC: ((("group AB" OR “Group A B” OR “Group A beta” or “Group A*”) NEAR strep*) or GABHS) 11311

#12 TITLE: streptococc* 58660

#13 (#9 or #10 or #11 or #12) 75,462

#14 (#8 AND #13) 3,938

#15 **(#4 OR #14)** 5,183

#16 TOPIC: (“clinical decision rule*” or “predicti* rule*” or “clinical predicti*”) 8052

#17 TITLE: predict* 786,816

#18 TOPIC: ((diagno* or classif* or discriminat* or differenti* or distinguish* or predict*) NEAR/3 (bacteri* or strep*)) 20943

#19 **(#16 or #17 or #18)** 810391

#20 ((diagnostic near/3 accuracy) OR (sensitivity AND specificity) OR "limit of detection" OR "reproducibility of result*" OR (diagnos* NEAR/3 differential*) OR “likelihood ratio” OR (predictive NEAR value*) OR ((accura* or reliab* or valid*) AND (bacteri* and (viral or virus*) AND (analys* or assay* or immunoassay* or classif* or detect* or diagnos* or differenti* or predict* or technique* or test*))) OR “observer variation*” OR “receiver operating characteristic*” OR “roc curve*” OR (false NEAR/2 (positiv* or negativ*))) 649,515

#21 TITLE: (sensitivity or specificity) 261560

#22 **(#20 or #21)** 899,345

#23 (#15 AND #19 AND #22) 160

#24 (#1 OR #23) 320

#### Ovid MEDLINE

<1946 to 24 June 2021>

1 feverpain*.mp. 10

2 McIsaac*.mp. 46

3 centor*.mp. 128

4 or/1-3 162

5 (strep* adj3 (throat? or tonsil* or pharyn* or nasopharyn* or epipharyn* or rhinopharyn*)).tw,kf. 3,400

6 GAS pharyngiti*.tw,kf. 214

7 (5 or 6) 3,467

8 pharyngitis/ or nasopharyngitis/ or retropharyngeal abscess/ or tonsillitis/ or peritonsillar abscess/ 16,048

9 pharyngiti*.tw,kf. 6,085

10 pharyngotonsilliti*.tw,kf. 389

11 (epipharyngiti* or nasopharyngiti* or rhinopharyngiti*).tw,kf. 1,281

12 pharyngalgi*.tw,kf. 100

13 ((throat? or tonsil* or pharyn* or nasopharyn* or epipharyng* or rhinopharyng*) adj3 (ache? or aching or infect* or pain* or sore*)).tw,kf. 11,363

14 (throat adj (culture? or swab?)).tw,kf. 3,855

15 or/8-14 30,262

16 bacterial infections/di 9,757

17 streptococcal infections/ 33,786

18 (group A beta h?emolytic strep* or group A B h?emolytic strep* or group AB h?emolytic strep* or ("group A" adj3 strep*) or GABHS).tw,kf. 10,205

19 (strep* adj3 infect*).tw,kf. 12,249

20 streptococc*.ti,kf. 57,697

21 or/16-20 84,823

22 (15 and 21) 5,709

23 (7 or 22) 6,629

24 clinical decision rules/ 603

25  (predicti* rule? or clinical* predicti*).tw. or predict*.ti,kf.  381,268

26  ((diagno* or classif* or discriminat* or differenti* or distinguish* or predict*) adj3 (bacteri* or strep*)).tw,kf.  16,785

27  or/24-26  397,206

28  (23 and 27)  504

29  "sensitivity and specificity"/ or "limit of detection"/ or roc curve/ or "predictive value of tests"/  602,753

30  "reproducibility of results"/  418,999

31  Diagnosis, Differential/  458,182

32  (sensitivity and specificity).tw. or (sensitivity or specificity).kf.  264,477

33  likelihood ratio.tw,kf.  12,692

34  (predictive adj4 value*).tw,kf.  120,484

35  ((accura* or reliab* or valid*) and (bacteri* and (viral or virus*) and (analys* or assay* or immunoassay* or classif* or detect* or diagnos* or differenti* or predict* or technique* or test*))).tw,kf.  2,759

36  observer variation*.tw,kf.  1,532

37  roc curve*.tw,kf.  40,063

38  receiver operating characteristic*.tw,kf.  81,123

39  (false adj (positiv* or negativ*)).tw,kf.  82,110

40  or/29-39  1,558,014

41  (28 and 40)  198

42  (4 or 41)  340

#### Ovid Embase

<1980 to 2021 Week 23>

1  feverpain*.mp. (19)
2   McIsaac*.mp. (70)
3  Centor*.mp. (198)
4  or/1-3 (257)
5  Streptococcal pharyngitis/ (1491)
6  (strep* adj3 (throat? or tonsil* or pharyn* or nasopharyn* or epipharyng* or rhinopharyng*)).tw,kw. (3838)
7  GAS pharyngiti*.tw,kw. (297)
8  or/5-7 (4637)
9   sore throat/ (18702)
10  pharyngitis/ (15257)
11  pharyngiti*.tw. (7285)
12  pharyngotonsilliti*.tw. (534)
13  (epipharyngiti* or nasopharyngiti* or rhinopharyngiti*).tw. (3017)
14   pharyngalgi*.tw. (122)
15  ((throat? or tonsil* or pharyn* or nasopharyn* or epipharyng* or rhinopharyng*) adj3 (ache? or aching or infect* or pain* or sore*)).tw. (15672)

16 (throat adj (culture? or swab?)).tw. (4768)
17 or/9-16 (47599)
18 bacterial infection/di [Diagnosis] (12686)
19 streptococcus infection/ (20349)
20 group a streptococcal infection/ (2265)
21 streptococcus group a/ (7183)
22 (group A beta h?emolytic strep* or group A B h?emolytic strep* or group AB h?emolytic strep* or ("group A" adj3 strep*) or GABHS).tw. (10423)
23 (strep* adj3 infect*).tw,kw. (11755)
24 streptococc*.ti,kw. (56235)
25 or/18-24 (86478)
26 8 or (17 and 25) (7756)
27 (predicti* rule? or clinical* predicti*).tw. or predict*.ti,kw. (547126)
28 ((diagno* or classif* or discriminat* or differenti* or distinguish* or predict*) adj3 (bacteri* or strep*)).tw,kw. (19330)
29 or/27-28 (565085)
30 26 and 29 (600)
31 diagnostic test accuracy study/ (154482)
32 diagnostic accuracy/ (266275)
33 differential diagnosis/ (334458)
34 "sensitivity and specificity"/ (395030)
35 predictive value/ (192432)
36 receiver operating characteristic/ (145448)
37 "limit of detection"/ (95335)
38 diagnostic error/ or false negative result/ or false positive result/ (103653)
39 (sensitivity and specificity).tw. or (sensitivity or specificity).kw. (392026)
40 likelihood ratio.tw,kw. (17696)
41 (predictive adj4 value*).tw,kw. (179921)
42 ((accura* or reliab* or valid*) and (bacteri* and (viral or virus*) and (analys* or assay* or immunoassay* or classif* or detect* or diagnos* or differenti* or predict* or technique* or test*))).tw,kw. (3861)
43 (observer adj variation*).tw,kw. (2332)
44 (roc adj curve*).tw,kw. (70451)
45 (false adj (positiv* or negativ*)).tw,kw. (109082)
46 or/31-45 (1487551)
47 30 and 46 (244)
48 4 or 47 (476)

### 2.0 Supplementary Table 1

**Supplementary table 1: Sensitivity and specificity of McIsaac and Centor scores from studies that provided 2x2 data compared to throat culture reference standard.** Summary estimates of sensitivity and specificity, false positive rate, positive and negative likelihood ratios for McIsaac and Centor scores at several thresholds. PPV and NPV calculated from a GAS prevalence of 24.1% in secondary care (6).*Results from sensitivity analysis using metandi in Stata, after noting unintuitively wide 95% CIs from the primary analysis. We were unable to do a repeat analysis using metandi for McIsaac score 0 and Centor score 0 due to an insufficient number of studies. PPV and NPV were calculated from summary 2x2 data for each score.

| Threshold | Sensitivity (95% CI) | Specificity (95% CI) | PPV | NPV | Likelihood Ratio +ve (95% CI) | Likelihood Ratio -ve (95% CI) |
|---|---|---|---|---|---|---|
| **McIsaac scores (n = 5 studies)** | | | | | | |
| ≥1 | 0.999 (0.505 – 1.0) *0.998 (0.859 - 1.000) | 0.001 (0 – 0.8) *0.00149 (0.000 - 0.384) | 0.24 | 0.70 | 0.999 (0.994 - 1.005) | 2.237 (0.008 - 627.441) |
| ≥2 | 0.959 (0.832 - 0.991) | 0.125 (0.044 - 0.306) | 0.26 | 0.91 | 1.095 (0.965 - 1.244) | 0.329 (0.082 - 1.321) |
| ≥3 | 0.819 (0.576 - 0.938) | 0.41 (0.242 - 0.603) | 0.31 | 0.88 | 1.389 (1.01 - 1.91) | 0.441 (0.174 - 1.12) |
| ≥4 | 0.478 (0.211 - 0.758) | 0.776 (0.625 - 0.878) | 0.40 | 0.82 | 2.133 (0.989 - 4.601) | 0.673 (0.377 - 1.2) |
| **Centor scores (n = 2 studies)** | | | | | | |
| ≥1 | 0.984 (0.894 - 0.998) | 0.052 (0.022 - 0.118) | 0.25 | 0.91 | 1.037 (0.981 - 1.097) | 0.313 (0.037 - 2.615) |
| ≥2 | 0.903 (0.801 - 0.956) | 0.237 (0.163 - 0.332) | 0.27 | 0.88 | 1.184 (1.032 - 1.359) | 0.408 (0.176 - 0.945) |
| ≥3 | 0.581 (0.455 - 0.696) | 0.516 (0.143 - 0.872) | 0.28 | 0.80 | 1.200 (0.450 - 3.204) | 0.812 (0.316 - 2.091) |
| ≥4 | 0.290 (0.191 - 0.414) | 0.853 (0.351 - 0.984) | 0.39 | 0.79 | 1.981 (0.252 - 15.593) | 0.832 (0.567 - 1.219) |

#### 3.0 Supplementary Figure 1

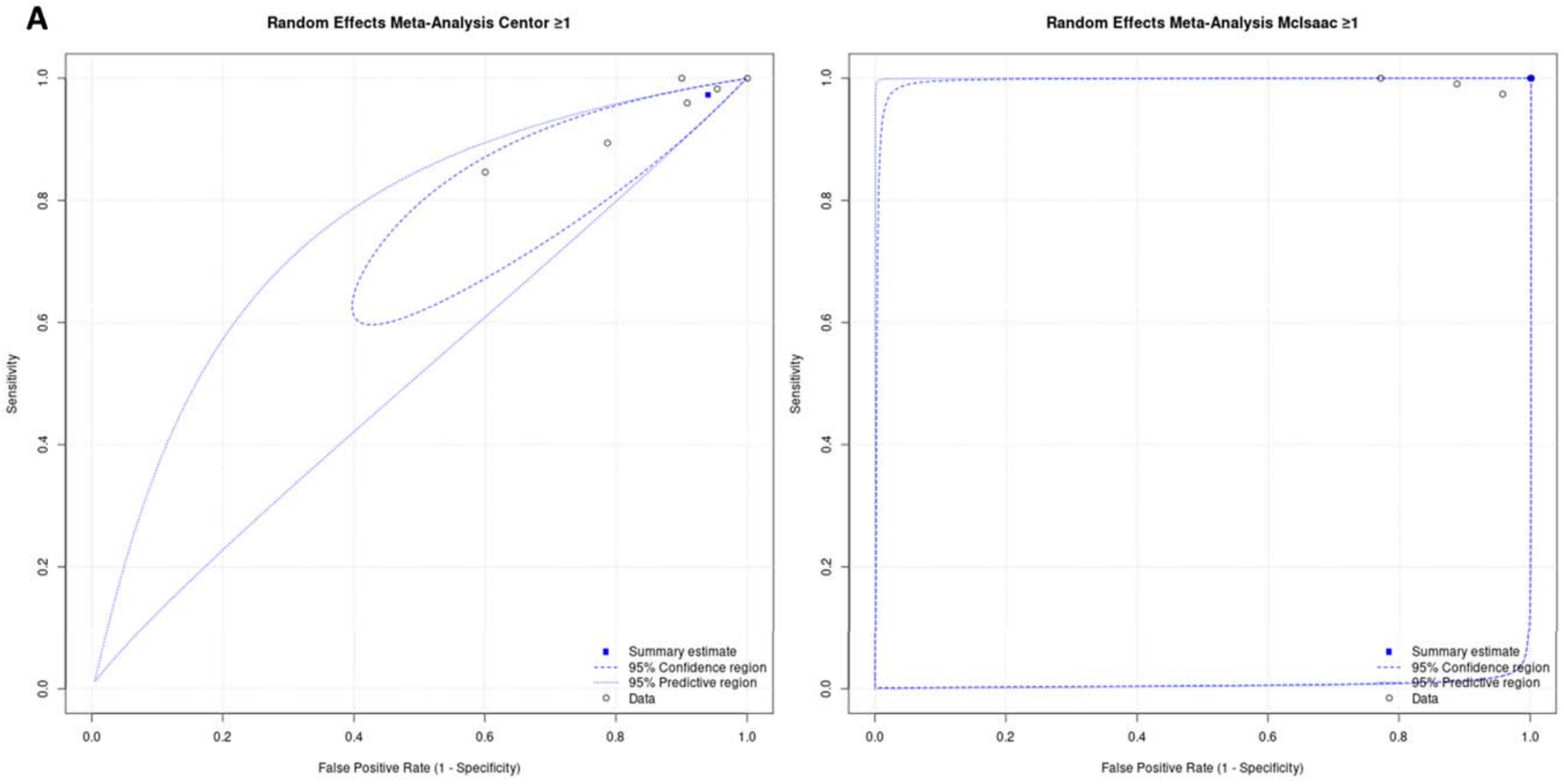

**B**

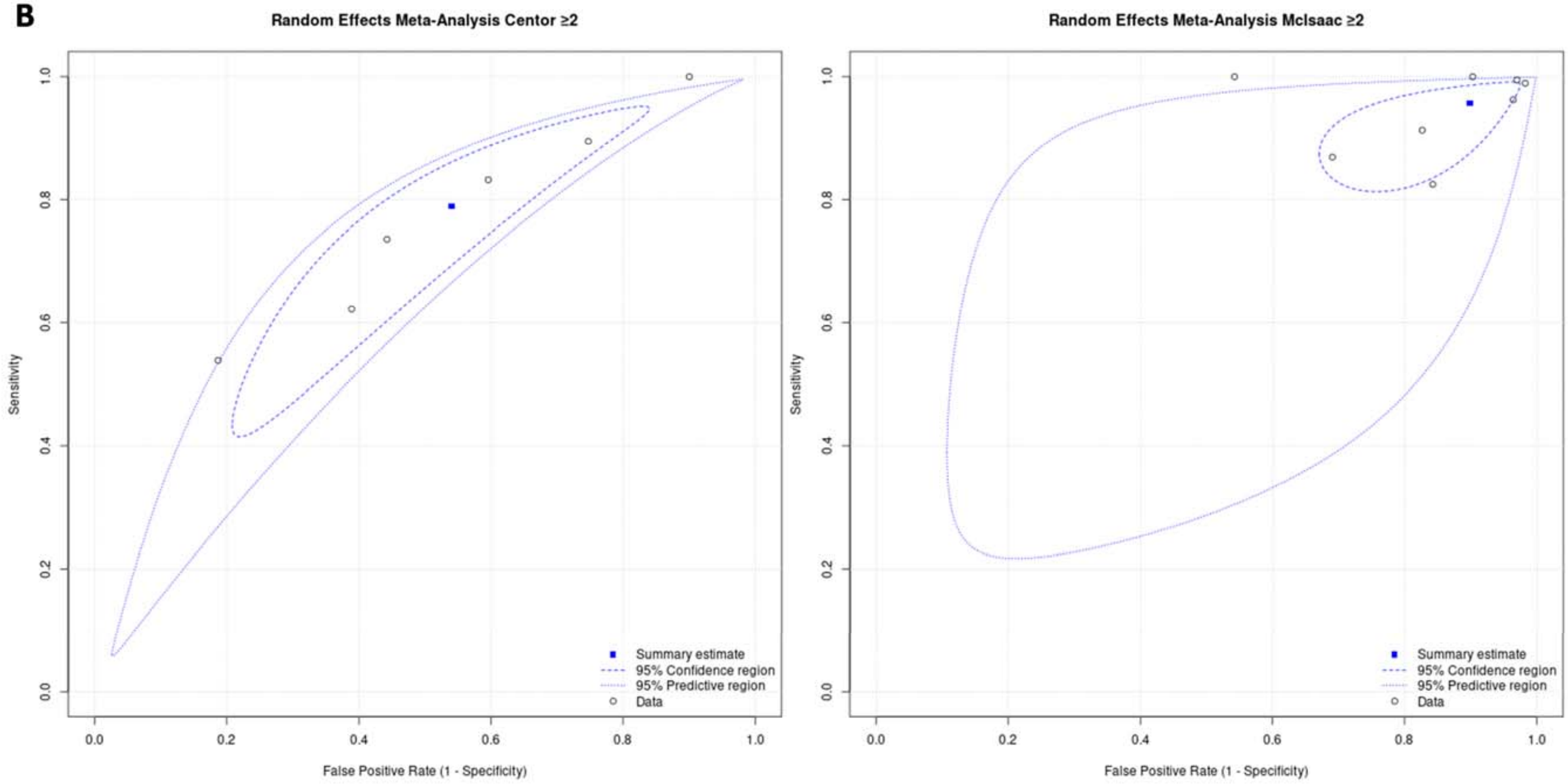
Random Effects Meta-Analysis Centor ≥2
Sensitivity
False Positive Rate (1 - Specificity)
Summary estimate
95% Confidence region
95% Predictive region
Data
Random Effects Meta-Analysis McIsaac ≥2
Sensitivity
False Positive Rate (1 - Specificity)
Summary estimate
95% Confidence region
95% Predictive region
Data

**C**

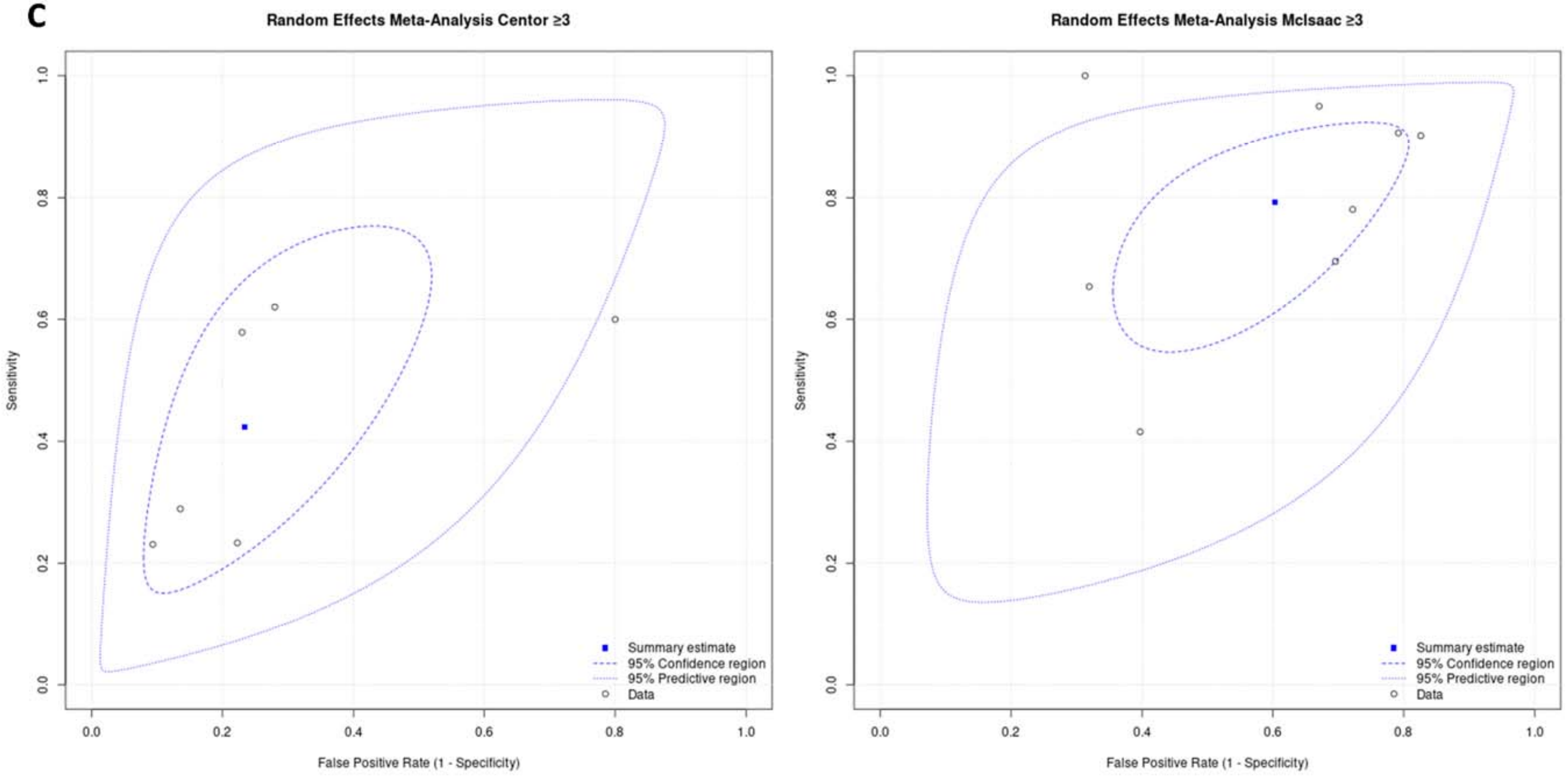

Random Effects Meta-Analysis Centor ≥3
Sensitivity
False Positive Rate (1 - Specificity)
Summary estimate
95% Confidence region
95% Predictive region
Data
Random Effects Meta-Analysis McIsaac ≥3
Sensitivity
False Positive Rate (1 - Specificity)
Summary estimate
95% Confidence region
95% Predictive region
Data

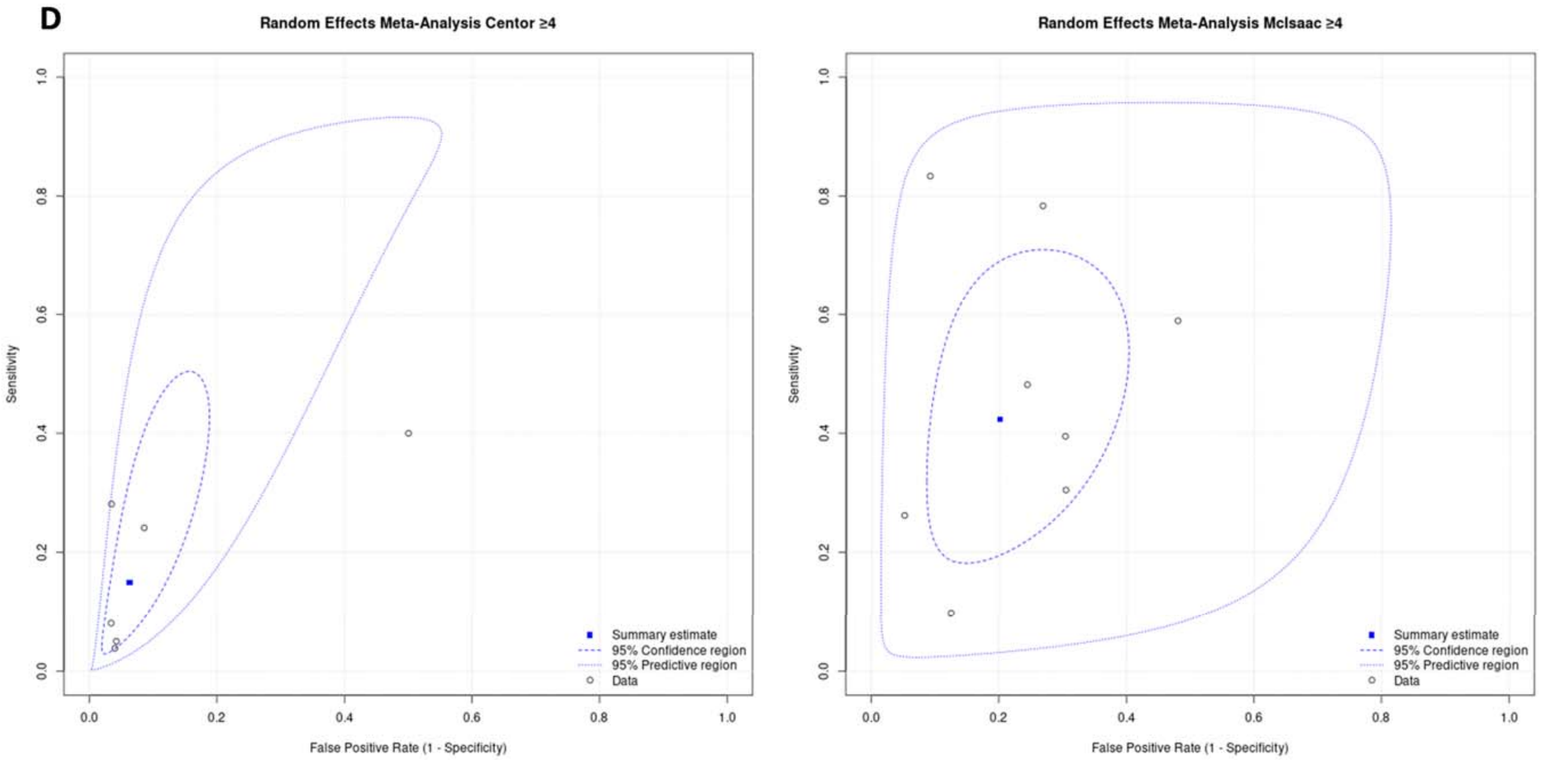


**Supplementary Figure 1A-D: SROC plots for Centor score (left) and McIsaac score (right) for thresholds 1-4.** Summary estimates shown with data points, SROC curve, and 95% confidence and predictive regions. SROC = summary ROC; ROC = receiver operating characteristic. 95% confidence region represents uncertainty about the summary estimates, whereas the 95% predictive region indicates the region that we can be 95% certain the sensitivity and specificity in a future study would lie, taking into account heterogeneity observed been estimates from previous studies.
